# The significance and characteristics of the patients with Ribbon-like signal hyperintensity after acute ischemic stroke

**DOI:** 10.1101/2023.08.21.23294396

**Authors:** Takeshi Miyamoto, Shu Sogabe, Masaaki Korai, Izumi Yamaguchi, Manabu Ishihara, Kenji Shimada, Yasuhisa Kanematsu, Kazutaka Kuroda, Yuki Yamamoto, Nobuaki Yamamoto, Masafumi Harada, Yuishin Izumi, Yasushi Takagi

## Abstract

**Background:** We discovered a novel finding of ribbon-like signal hyperintensity of the cerebral cortical gyri, named the ribbon sign, after reperfusion therapy. Herein, we report the significance and clinical characteristics of ribbon signs.

**Methods:** Data from consecutive patients with acute ischemic stroke and anterior large-vessel occlusion were prospectively extracted from the Tokushima University Hospital Stroke Registry between January 2011 and March 2020. Diffusion-weighted imaging (DWI) was retrospectively assessed in patients with acute ischemic stroke with large-vessel occlusion, with or without treatment.

**Results:** A total of 140 patients (78 males, Average age: 75.7 years) were enrolled in the study. The mean DWI-Alberta Stroke Program Early Computed Tomography Score (DWI- ASPECTS) was 7.0. Among the patients, 113 (80.7%) underwent reperfusion therapy and 95 (67.9%) had unfavorable outcomes. Eighty-one (57.9%) patients underwent successful recanalization. The ribbon sign was more common in patients with successful recanalization than in the patients with unsuccessful recanalization (53.1% vs. 8.5%, respectively; *p*<0.01).

**Conclusion:** Our study is the first to report that the ribbon sign is a specific finding after successful recanalization in patients with acute ischemic stroke.

## INTRODUCTION

The efficacy and safety of reperfusion therapy, including intravenous thrombolysis (IVT) and endovascular treatment (EVT), in patients with acute ischemic stroke (AIS) due to anterior large-vessel occlusion (LVO) have been established (1–8). A non-invasive intracranial vascular study is recommended during the initial imaging evaluation of patients with acute stroke. Diffusion-weighted imaging (DWI) lesions at onset have been accepted as the ischemic core in some clinical studies. However, whether DWI lesions at onset are already infarcted or potentially salvageable is debatable, particularly in cases where rapid reperfusion is possible.

We assessed magnetic resonance imaging (MRI) as first-line and post-treatment imaging in patients with acute ischemic stroke and discovered a novel finding of ribbon-like signal hyperintensity of the cerebral cortical gyri, named the ribbon sign, after reperfusion therapy. Herein, we report the significance and clinical characteristics of ribbon signs after reperfusion therapy.

## METHODS

Data from consecutive patients with AIS caused by anterior LVO were prospectively extracted from the Tokushima University Hospital Stroke Registry between January 2011 and March 2020. MRI was implemented as the first-line pretreatment imaging. Patients who fulfilled the following eligibility criteria were included: (1) imaging both within 24 hours of AIS onset and within 72 hours posttreatment; (2) baseline MR angiography to confirm an internal carotid artery (ICA) or middle cerebral artery (MCA) M1 (horizontal segment of MCA) occlusion; and AIS confirmed by baseline MRI (3T; DWI: *b*=0–1000 s/mm^2^:5-mm contiguous slices); and (3) patients with clinical outcome obtained at 3 months after onset.

Clinical characteristics including baseline demographics, baseline National Institutes of Health Stroke Scale score (NIHSS), onset to picture, lesion side, baseline DWI-Alberta Stroke Program Early Computed Tomography Score (DWI-ASPECTS), stroke subtype, type of reperfusion therapy, outcomes, and ribbon sign were retrospectively collected. The ribbon sign was defined as ribbon-like signal hyperintensity of the cerebral cortical gyri on DWI on the second MR within 72 h post-treatment.

If eligible, patients received IVT (0.6 mg/kg alteplase) according to the Japanese-approved standard care protocol before EVT. All interventional procedures were performed by board-certified neurointerventionalists of the Japanese Society for Neuroendovascular Therapy. Successful reperfusion was defined as modified Thrombolysis in Cerebral Infarction Scale grade 2b or 3 after EVT and a decrease of 4 points in the NIHSS score after IVT or medical therapy. Recanalization was defined as successful reperfusion and patency of the ipsilateral internal carotid artery and middle cerebral artery on the second MR angiography within 72 h post-treatment.

Two stroke neurologists (Y.Y. and N.Y.) independently evaluated baseline DWI- ASPECTS on MRI at onset and the ribbon sign on MRI within 72 h post-treatment. When the readers’ assessments differed, a final consensus was reached by a third reader, a neuroradiologist (M.Y.).

The study protocol was approved by the Tokushima University Hospital Ethics Committee. Written informed consent was obtained from the patients.

### Statistical analysis

Continuous variables were described as mean±standard deviation (SD) and compared using Student’s t-test. Categorical variables were compared using χ^2^ or Fisher’s exact test as appropriate. Differences were considered statistically significant at *p*<0.05. All statistical analyses were performed using GraphPad Prism version 9 (GraphPad Software, California, USA).

## RESULTS

### Patients

During the study period, 221 patients were diagnosed with AIS due to anterior LVO. Among these patients, 70 (31.7%) were excluded due to the lack of MRI findings within 72 hours post-treatment. Among these, 11 patients were excluded due to missing outcomes three months after onset. Finally, a total of 140 patients were included in this analysis. The baseline clinical characteristics of the patients are presented in Table 1. The etiology of ischemic stroke was diagnosed as intracranial atherosclerotic disease or embolic stroke due to a cardiac source, another origin, or an unknown origin. A total of 113 (80.7%) patients underwent reperfusion therapy (intravenous thrombolysis [IVT], n=21 [15.0%]; endovascular treatment [EVT], n=42 [30.0%]; combined IVT/EVT, n=50 [35.7%]). Recanalization was achieved in 81 (57.9%) patients. A total of 45 (32.1%) patients had a modified Rankin Scale (mRS) 0–2 and 95 (67.9%) had an mRS 3–6 at three months after onset. The ribbon sign was observed in 48 (34.3%) patients at 2^nd^ MRI.

**Table 1.**
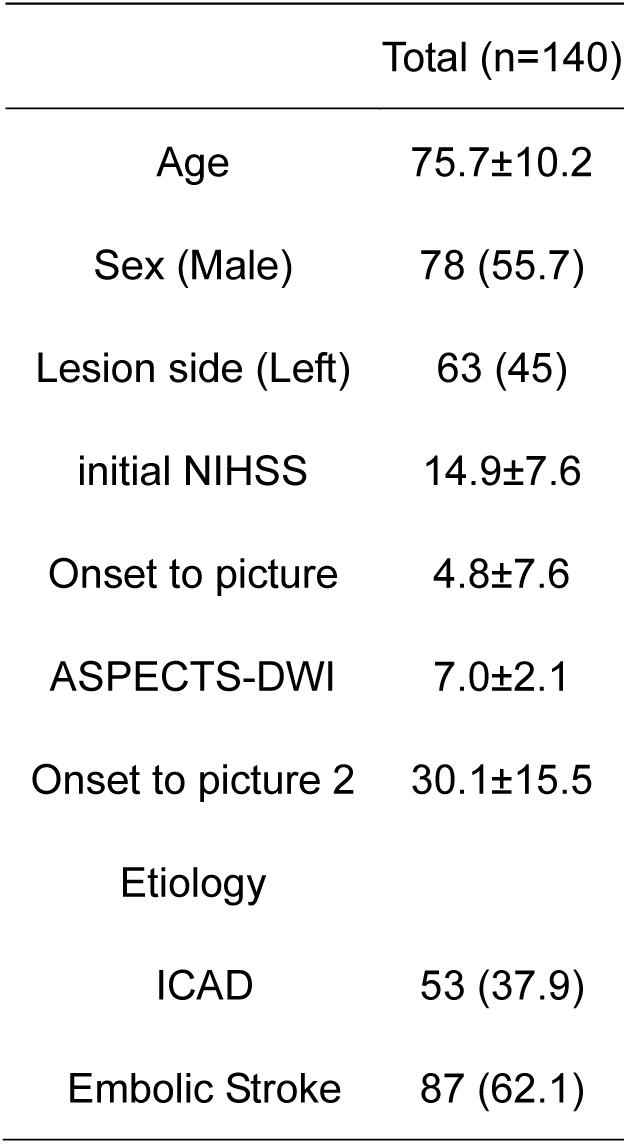
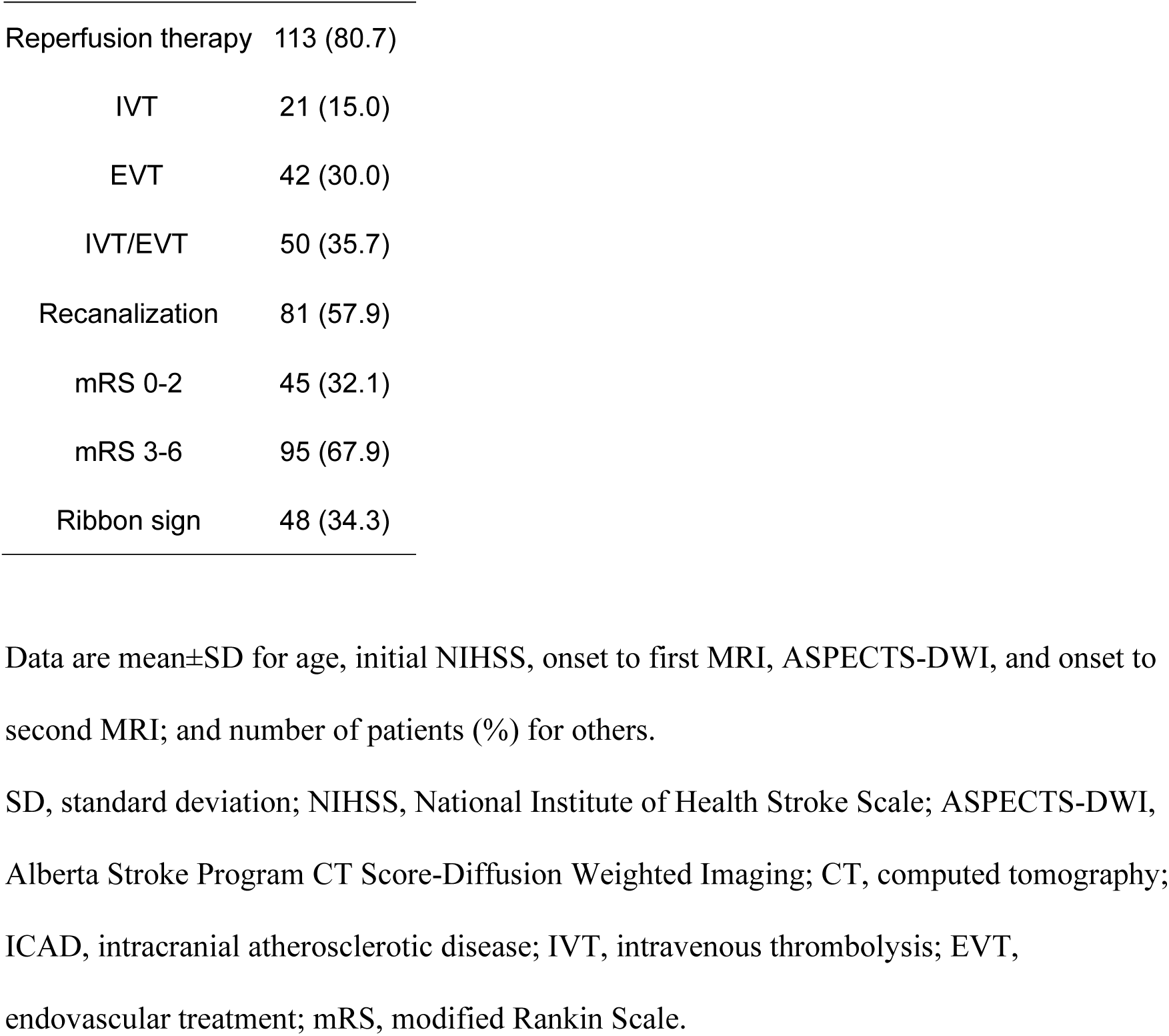
Characteristics of the patients in this study.

### Illustrative Cases

Representative cases of acute ischemia with anterior LVO are shown in Figures 1 and 2. Both were diagnosed with acute ischemic stroke and proximal middle cerebral artery occlusion.

**Figure 1.**
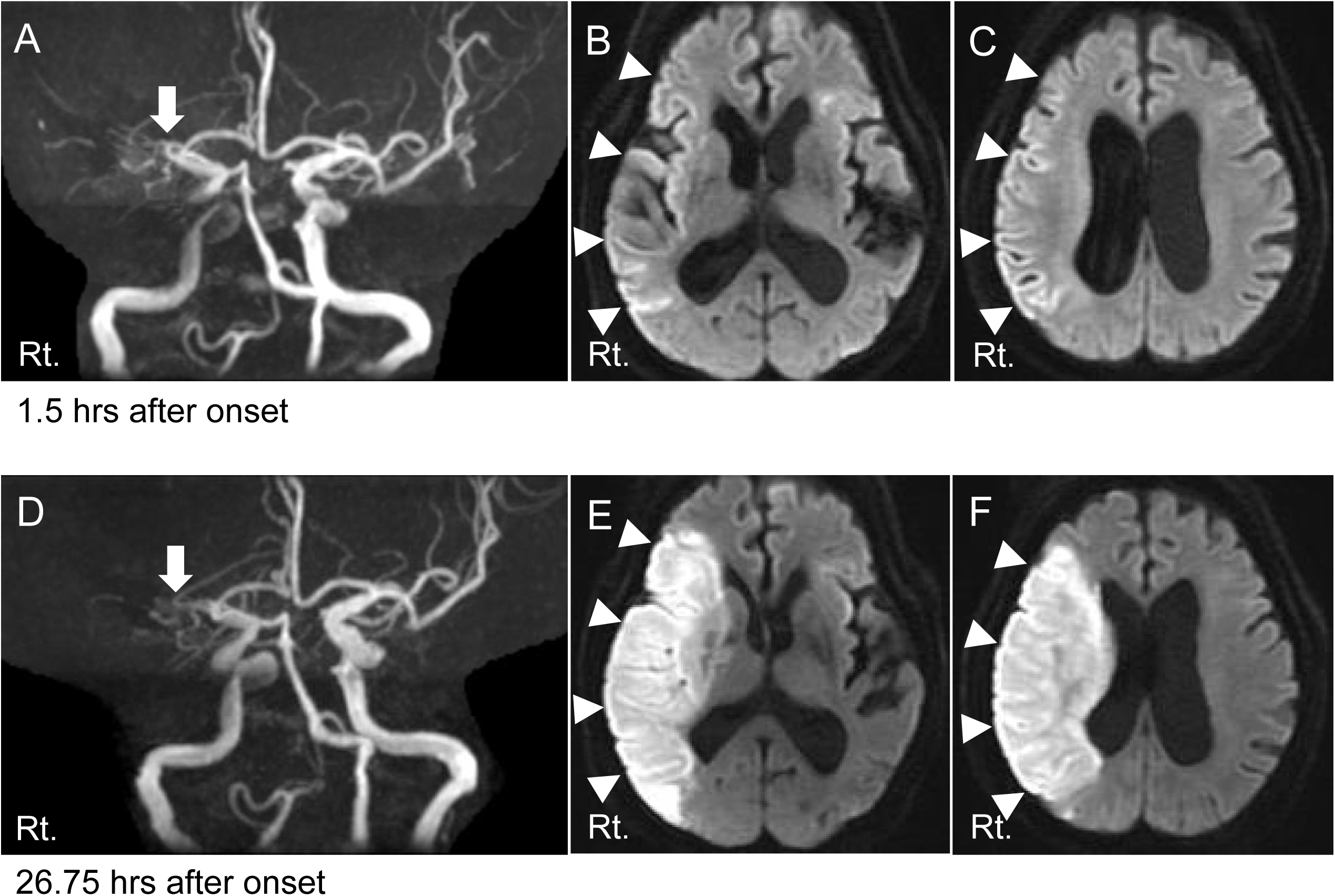
A: MRA showing occlusion of right middle cerebral artery (MCA) at the stem 1.5 hours after onset (arrow). B, C: DWI showing slight hyperintensity in the right MCA territory (arrowhead). D: MRA shows occlusion of the right MCA at the stem 26.75 hours after onset (arrow). DWI showing bright hyperintensity in the right MCA territory without recanalization of the MCA (arrowhead). MRA, indicates Magnetic Resonance Angiography; DWI, Diffusion Weighted Imaging.

**Figure 2.**
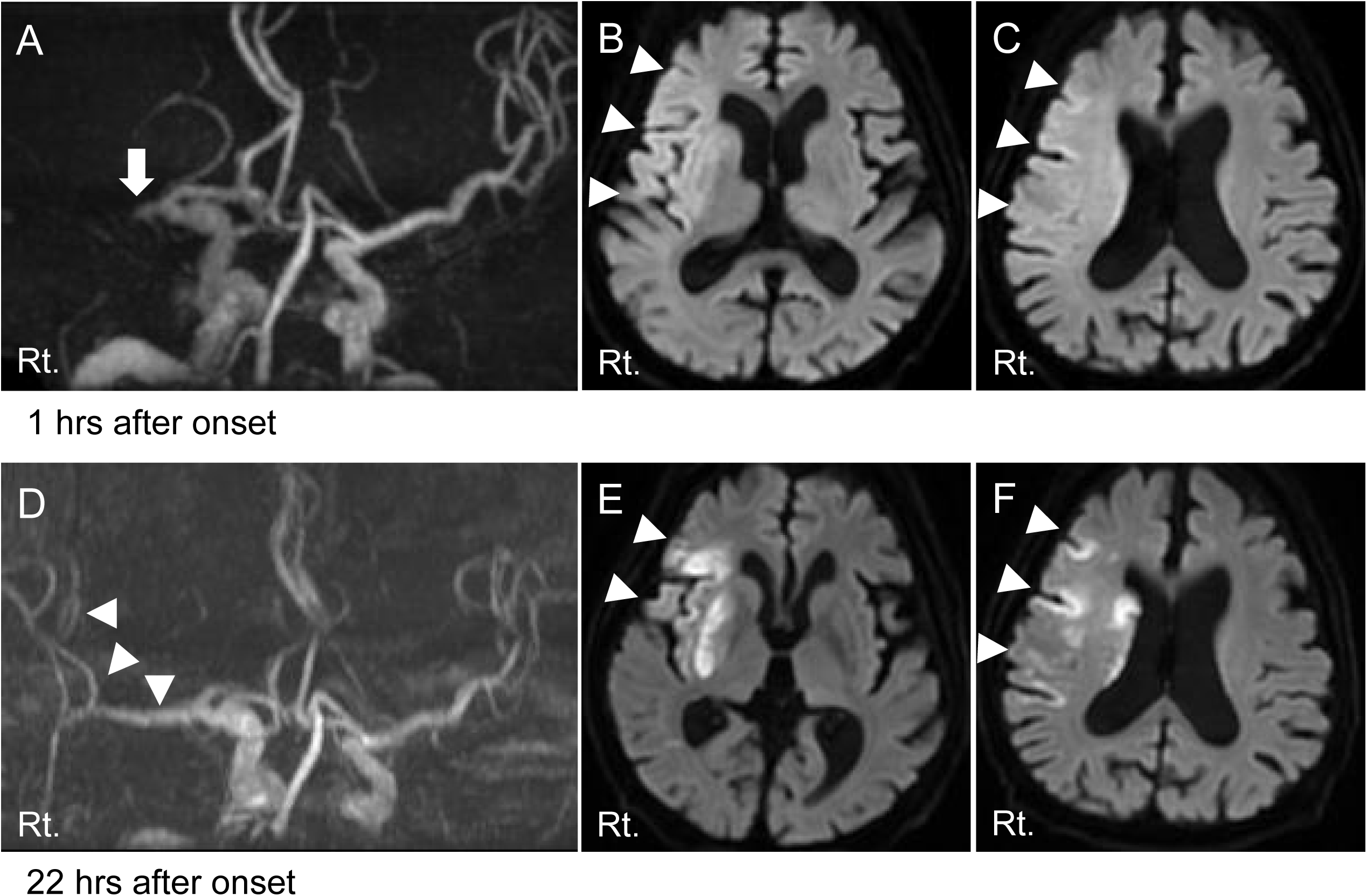
(**A**) MRA shows occlusion of right middle cerebral artery (MCA) at the stem 1.0 hour after onset (arrow). (**B, C**) DWI shows a slight hyperintensity at the right MCA territory (arrow head). (**D**) MRA shows recanalization of right MCA at the stem 22 hours after onset (arrow head). (**E, F**) DWI shows a bright hyperintensity at the cortical gyri and basal ganglia of right MCA territory with recanalization of MCA (arrow head). MRA, Magnetic Resonance Angiography; DWI, Diffusion Weighted Imaging.

#### Case 1

In case 1, the patient was treated conservatively without reperfusion therapy because of extensive ischemic lesions on DWI (DWI-ASPECTS=2) at 1.5 hours after onset. The second DWI showed a massive ischemic lesion in both the cortex and white matter 27 h after onset. Three months after onset, the patient had an mRS score of 4.

#### Case 2

In Case 2, the patient underwent reperfusion therapy with combined IVT and EVT. Although extensive ischemic lesions were observed on the first DWI (DWI-ASPECTS=4) 1 h after onset, 2^nd^ MRI at 22 h after onset showed a DWI lesion limited to the cerebral cortical gyri and basal ganglia, which is called the Ribbon sign. Three months after onset, the patient had an mRS score of 3.

### Ribbon Signs Were Observed in the Patients with Acute Ischemic Stroke after Recanalization

We divided the patients with anterior LVO into two groups based on recanalization status after treatment (Table 2). The patients with recanalization had left-side lesions (58.0% vs 32.2%, respectively, *p*=0.0035), higher initial NIHSS scores (16.1±8.1 vs 13.1±6.7, respectively, *p*=0.0214), earlier evaluation of first MRI after onset (3.0±34 vs 7.3±10.5, respectively, *p*=0.0008) and earlier evaluation of second MRI after treatment (27.9±12.9 vs 33.1±18.0, respectively, *p*=0.0451). Among the patients with recanalization, 77 (95.1 %) received reperfusion therapy (IVT, n=14; EVT, n=24; combined IVT/EVT, n=39). Four (4.9%) experienced spontaneous recanalization. The patients who underwent recanalization had more favorable outcomes at 3 months after onset (37 [45.7%] vs. 8 [13.6%], *p*<0.0001) and a higher incidence of the ribbon sign (43 [53.1%] vs. 5 [8.5%], *p*<0.0001).

**Table 2.** Characteristics of the patients with or without recanalization.

|  | Recanalization - (n=59) | Recanalization + (n=81) | P value |
| --- | --- | --- | --- |
| Age | 74.0±10.7 | 77.0±9.8 | 0.0894 |
| Sex (Male) | 33 (56.0) | 45 (55.6) | >0.9999 |
| Lesion side (Left) | 19 (32.2) | **47 (58.0)** | **0.0035** |
| initial NIHSS | 13.1±6.7 | **16.1±8.0** | **0.0214** |
| Onset to first MRI | 7.3±10.5 | **3.0±3.4** | **0.0008** |
| ASPECTS-DWI | 7.0±2.4 | 7.0±1.9 | 0.8834 |
| Onset to second MRI | 33.1±18.0 | 27.9±12.9 | **0.0451** |
| Etiology |  |  | 0.0535 |
| ICAD | 28 (47.5) | 25 (30.9) |  |
| Embololic Stroke | 31 (52.5) | 56 (69.1) |  |
| Reperfusion therapy | 36 (61.0) | 77 (95.1) | **<0.0001** |
| IVT | 7 (11.9) | 14 (17.3) |  |
| EVT | 18 (30.5) | 24 (29.6) |  |
| IVT/EVT | 11 (18.6) | 39 (48.1) |  |
| mRS 0-2 | 8 (13.6) | 37 (45.7) |  |
| mRS 3-6 | 51 (86.4) | **44 (54.3)** | **<0.0001** |
| Ribbon sign | **5 (8.5)** | **43 (53.1)** | **<0.0001** |

Data are mean±SD for age, initial NIHSS, onset to first MRI, ASPECTS-DWI, and onset to second MRI; and number of patients (%) for others.
SD, standard deviation; NIHSS, National Institute of Health Stroke Scale; ASPECTS-DWI, Alberta Stroke Program CT Score-Diffusion Weighted Imaging; CT, computed tomography; ICAD, intracranial atherosclerotic disease; IVT, intravenous thrombolysis; EVT, endovascular treatment; mRS, modified Rankin Scale.

### Ribbon Sign Was Associated with Lower DWI-ASPECTS

To analyze the characteristics of the ribbon sign, we divided the patients with recanalization into two groups according to the status of the ribbon sign (Table 3). The patients with ribbon signs had lower DWI-ASPECTS (6.0±1.6 vs 8.2±1.6, respectively). Despite the low ASPECTS, the rates of reperfusion therapy (93% vs. 97.4%, *p*=0.6184) and favorable outcomes (44.2% vs. 44.7%, *p*>0.9999) did not differ between the two groups.

**Table 3.** Characteristics of the patients with recanalization with or without Ribbon sign.

|  | Ribbon – (n=38) | Ribbon + (n=43) | P value |
| --- | --- | --- | --- |
| Age | 77.6±8.6 | 76.4±10.8 | 0.5985 |
| Sex (Male) | 23 (60.5) | 22 (51.2) | 0.5024 |
| Lesion side (Left) | 22 (57.9) | 25 (58.1) | >0.9999 |
| initial NIHSS | 15.6±7.1 | 16.6±8.8 | 0.5683 |
| Onset to first MRI | 3.1±3.7 | 2.8±3.2 | 0.7321 |
| ASPECTS-DWI | 8.2±1.6 | **6.0±1.6** | **<0.0001** |
| Cortex-DWI positive | 16 (42.1) | **42 (97.7)** | **<0.0001** |
| Onset to second MRI | 28.8±14.6 | 26.9±11.2 | 0.5175 |
| Etiology |  |  | 0.8116 |
| ICAD | 11 (28.9) | 14 (32.6) |  |
| Embollic Stroke | 27 (71.1) | 29 (67.4) |  |
| Reperfusion therapy | 37 (97.4) | 40 (93.0) | 0.6184 |
| IVT | 4 (10.5) | 10 (23.3) |  |
| EVT | 14 (36.8) | 10 (23.3) |  |
| IVT/EVT | 19 (50.0) | 20 (46.5) |  |
| mRS 0-2 | 17 (44.7) | 19 (44.2) | >0.9999 |
| mRS 3-6 | 21 (58.2) | 24 (55.8) |  |

Data are mean±SD for age, initial NIHSS, onset to first MRI, ASPECTS-DWI, and onset to second MRI; and number of patients (%) for others.
SD, standard deviation; NIHSS, National Institute of Health Stroke Scale; ASPECTS-DWI,
Alberta Stroke Program CT Score-Diffusion Weighted Imaging; CT, computed tomography;
ICAD, intracranial atherosclerotic disease; IVT, intravenous thrombolysis; EVT,
endovascular treatment; mRS, modified Rankin Scale.

### Ribbon Sign Was Observed in the Patients with Low ASPECTS after Recanalization

Patients with low ASPECTS were selected and divided into two groups based on the recanalization status after treatment (Table 4). The patients with recanalization had higher ASPECTS (4.5±0.81 vs 3.5±1.3, *p*=0.0132). Sixteen (84.2%) patients with recanalization underwent reperfusion therapy (IVT, n=2; EVT, n=5; combined IVT/EVT, n=9). The patients with recanalization had a more favorable outcome (mRS 0–3) at 3 months after onset (9 [47.4%] vs. 1 [7.1%], *p*=0.0209) and a higher incidence of the ribbon sign (16 [84.2%] vs. 1 [7.1%], *p*<0.0001).

**Table 4.** Characteristics of the patients with low ASPECTS with or without Recanalization.

|  | Recanalization - (n=14) | Recanalization + (n=19) | P value |
| --- | --- | --- | --- |
| Age | 71.8±13.1 | 75.9±11.7 | 0.0894 |
| Sex (Male) | 10(71.4) | 10(52.6) | 0.3441 |
| Lesion side (Left) | 3 (21.4) | 8 (42.1) | 0.2783 |
| initial NIHSS | 15.9±4.5 | 18.4±8.0 | 0.3048 |
| Onset to first MRI | 5.9±7.1 | 3.4±3.4 | 0.1900 |
| ASPECTS-DWI | 3.5±1.3 | 4.5±0.8 | **0.0132** |
| Onset to second MRI | 29.0±18.0 | 28.6±8.9 | 0.1287 |
| Etiology |  |  | >0.9999 |
| ICAD | 3 (21.4) | 5 (26.3) |  |
| Embollic Stroke | 11 (78.6) | 14 (73.7) |  |
| Reperfusion therapy | 10 (71.4) | 16 (84.2) | **0.0035** |
| IVT | 2 (14.3) | 2 (10.5) |  |
| EVT | 2 (14.3) | 5 (26.3) |  |
| IVT/EVT | 0 (0) | 9 (47.4) |  |
| mRS 0-3 | 1 (7.1) | 9 (47.4) |  |
| mRS 4-6 | 13 (92.9) | **10 (52.6)** | **0.0209** |
| Ribbon sign | **1 (7.1)** | **16 (84.2)** | **<0.0001** |

Data are mean±SD for age, initial NIHSS, onset to first MRI, ASPECTS-DWI, and onset to second MRI; and number of patients (%) for others.
SD, standard deviation; NIHSS, National Institute of Health Stroke Scale; ASPECTS-DWI,
Alberta Stroke Program CT Score-Diffusion Weighted Imaging; CT, computed tomography;
ICAD, intracranial atherosclerotic disease; IVT, intravenous thrombolysis; EVT,
endovascular treatment; mRS, modified Rankin Scale.

### Ribbon Sign Tended to Be Associated with Reperfusion Therapy and Favorable Outcome in the Patients with Low ASPECTS

To analyze the characteristics of the ribbon sign in patients with low ASPECTS, we divided patients with recanalization into two groups based on the status of the ribbon sign (Table 5). The patients with ribbon sign had higher DWI-ASPECTS (4.5±0.8 vs 3.6±1.3). The rate of reperfusion therapy tended to be higher in the patients with ribbon sign (82.4% vs. 37.5%, *p*=0.0501). The patients with ribbon sign had favorable outcomes at discharge (41.2% vs. 6.3%, respectively, *p*=0.0391) and at 3 months (47.1% vs. 12.5%, *p*=0.0570).

**Table 5.**
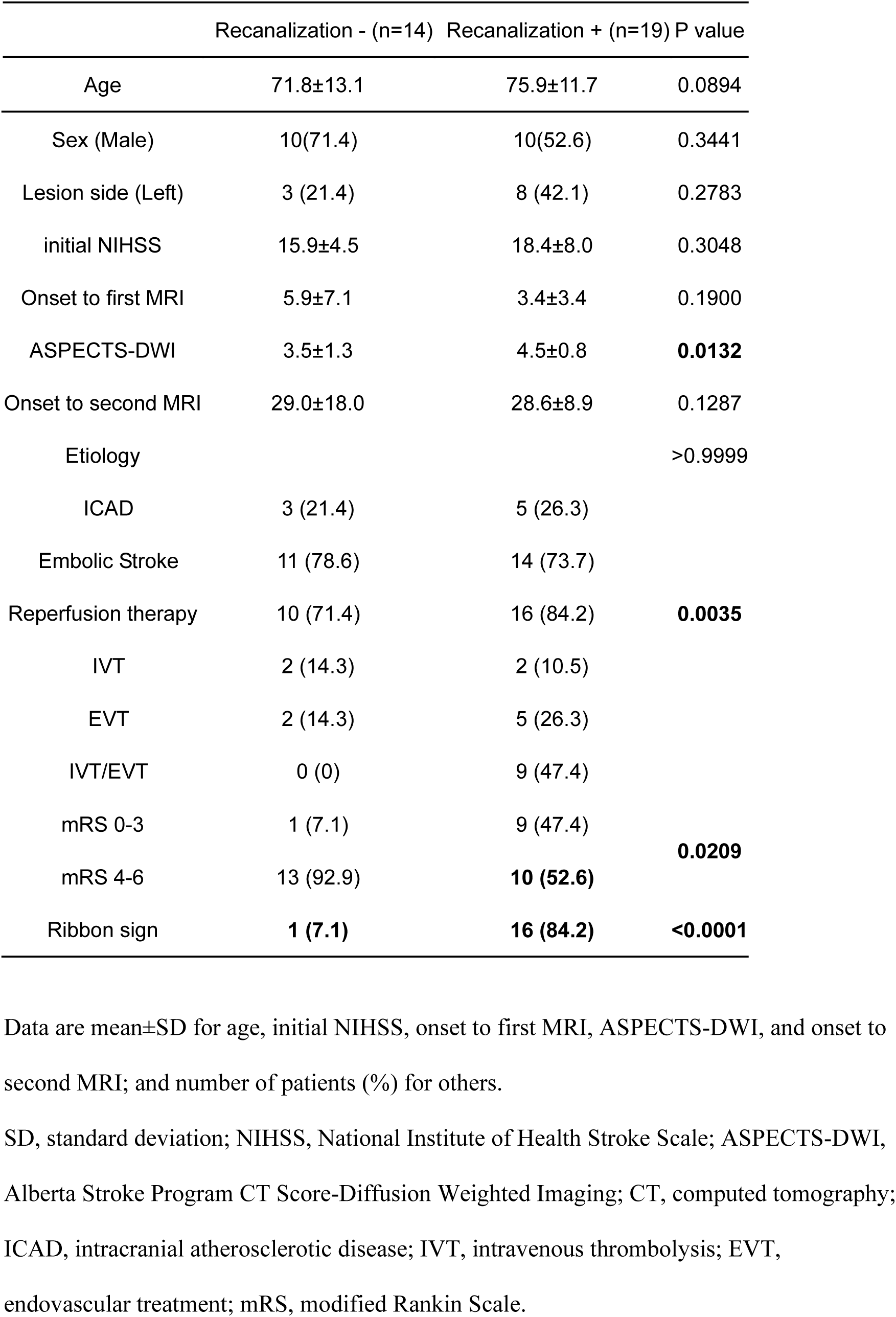
Characteristics of the patients with low ASPECTS with or without Ribbon sign.

|  | Recanalization - (n=14) | Recanalization + (n=19) | P value |
| --- | --- | --- | --- |
| Age | 71.8±13.1 | 75.9±11.7 | 0.0894 |
| Sex (Male) | 10(71.4) | 10(52.6) | 0.3441 |
| Lesion side (Left) | 3 (21.4) | 8 (42.1) | 0.2783 |
| initial NIHSS | 15.9±4.5 | 18.4±8.0 | 0.3048 |
| Onset to first MRI | 5.9±7.1 | 3.4±3.4 | 0.1900 |
| ASPECTS-DWI | 3.5±1.3 | 4.5±0.8 | **0.0132** |
| Onset to second MRI | 29.0±18.0 | 28.6±8.9 | 0.1287 |
| Etiology |  |  | >0.9999 |
| ICAD | 3 (21.4) | 5 (26.3) |  |
| Embollic Stroke | 11 (78.6) | 14 (73.7) |  |
| Reperfusion therapy | 10 (71.4) | 16 (84.2) | **0.0035** |
| IVT | 2 (14.3) | 2 (10.5) |  |
| EVT | 2 (14.3) | 5 (26.3) |  |
| IVT/EVT | 0 (0) | 9 (47.4) |  |
| mRS 0-3 | 1 (7.1) | 9 (47.4) | **0.0209** |
| mRS 4-6 | 13 (92.9) | **10 (52.6)** |  |
| Ribbon sign | **1 (7.1)** | **16 (84.2)** | **<0.0001** |

Data are mean±SD for age, initial NIHSS, onset to first MRI, ASPECTS-DWI, and onset to second MRI; and number of patients (%) for others.
SD, standard deviation; NIHSS, National Institute of Health Stroke Scale; ASPECTS-DWI,
Alberta Stroke Program CT Score-Diffusion Weighted Imaging; CT, computed tomography;
ICAD, intracranial atherosclerotic disease; IVT, intravenous thrombolysis; EVT,
endovascular treatment; mRS, modified Rankin Scale.

## DISCUSSION

The effectiveness of reperfusion therapy for anterior LVO is well established (1–8). We implemented MRI as the first-line pretreatment imaging. Using MRI, we demonstrated several findings of acute ischemic stroke (10,11). In the era of mechanical thrombectomy, most patients receiving reperfusion therapy achieve successful recanalization. We noticed that some patients who received reperfusion therapy had a ribbon-like signal hyperintensity in the cerebral cortical gyri on DWI after successful recanalization. We named this the ribbon sign and retrospectively analyzed the incidence and characteristics of acute ischemic stroke. Among the patients with successful recanalization, 53.1% had the ribbon sign. Only 8.5% of the patients without recanalization had a ribbon sign. Therefore, the ribbon sign is a characteristic finding in patients with acute ischemic stroke and anterior LVO after successful recanalization.

Next, we analyzed patients with successful recanalization and divided them into two groups: with or without the ribbon sign. The patients with Ribbon sign had lower ASPECTS- DWI score (6.0±1.6 vs 8.2±1.6, respectively) and a higher rate of DWI positivity in the cortex (97.7% vs 42.1%). In other words, patients with extensive DWI lesions, including the cortex, exhibited the ribbon sign after successful recanalization. Due to the differences in patient characteristics before reperfusion therapy, there was no difference in functional outcomes between the two groups. The ribbon sign is limited to the cortex, which may indicate DWI reversal of the white matter after reperfusion therapy. In patients with low ASPECTS, the ribbon sign tended to be associated with reperfusion therapy and favorable outcomes.

DWI is widely used to evaluate ischemic core (12,13). The sub-analysis of DEFUSE 2 demonstrated that the region of DWI reversal is transient and results in abnormalities in FLAIR. However, whether DWI precisely reveals the ischemic core is controversial (9,15,16). Reversible acute DWI lesions are found in half of the patients treated with intravenous tissue-type plasminogen activator (tPA) within 4.5 hours and DWI reversal is associated with good clinical outcomes. Gray matter and white matter may have differential vulnerabilities to acute ischemia. A previous study demonstrated that white matter is more prone to DWI reversal than gray matter. Consistent with this finding, the ribbon sign, which is limited to the gray matter, may indicate a DWI reversal in the white matter.

In the era of EVT, patients with LVO experience faster reperfusion than those treated with IVT alone. In animal studies, DWI reversal indicated limited necrosis of the brain tissue. Therefore, the initial DWI lesion may contain viable brain tissue. The ribbon sign may indicate incomplete infarction after revascularization therapy. Further studies are needed to elucidate the clinical significance of the ribbon sign and its relationship with clinical outcomes.

Our study has a limitation. This is a retrospective study. The patients with low ASPECTS may not treated with EVT. It have biased our results toward favorable outcome and caused some overestimation of the patients with low ASCPECTS after recanalization.

Our study is the first to report that the ribbon sign is a specific finding after successful recanalization in acute ischemic stroke patients. The ribbon sign appeared frequently in patients with large DWI lesions, including cortex, after recanalization.

## Data Availability

Raw data were generated at Tokushima University Hospital. Derived data supporting the findings of this study are available from the corresponding author [TM] on request.

## Non-standard Abbreviations and Acronyms

DWI: Diffusion-weighted imaging
DWI-ASPECTS: DWI-Alberta Stroke Program Early Computed Tomography Score
IVT: intravenous thrombolysis
EVT: endovascular treatment
AIS: acute ischemic stroke
LVO: large-vessel occlusion
MRI: magnetic resonance imaging
ICA: internal carotid artery
MCA: middle cerebral artery
NIHSS: National Institutes of Health Stroke Scale score
SD: standard deviation
mRS: modified Rankin Scale
tPA: tissue-type plasminogen activator

## Acknowledgments

We thank Drs Miyamoto and Sogabe performed in drafting and critical revision of the article. The remaining authors participated in critical revision of the article.

## Sources of funding

This work was supported by a Grant-in-Aid for Young Scientists (JSPS KAKENHI Grant Number 20K17933).

## Disclosure

None.

## Authors’ contributions

TM participated in the conception, design, data acquisition, and drafting of the article. SS, MK, IY, MI, KS, YK, KK, YY, and Y. Y. participated in data acquisition, analysis, and interpretation. MH, YI and YT critically reviewed the manuscript for intellectual content and approved its submission for publication.

## Notes

### Competing Interest Statement

The authors have declared no competing interest.

## References

1. Albers GW, Marks MP, Kemp S, Christensen S, Tsai JP, Ortega-Gutierrez S, McTaggart RA, Torbey MT, Kim-Tenser M, Leslie-Mazwi T, et al. Thrombectomy for stroke at 6 to 16 hours with selection by perfusion imaging. New England Journal of Medicine 2018;378:708–718.

2. Berkhemer OA, Fransen PS, Beumer D, van den Berg LA, Lingsma HF, Yoo AJ, Schonewille WJ, Vos JA, Nederkoorn PJ, Wermer MJ, et al. A randomized trial of intraarterial treatment for acute ischemic stroke. New England Journal of Medicine 2015;372:11–20.

3. Campbell BC, Mitchell PJ, Kleinig TJ, Dewey HM, Churilov L, Yassi N, Yan B, Dowling RJ, Parsons MW, Oxley TJ, et al. Endovascular therapy for ischemic stroke with perfusion-imaging selection. New England Journal of Medicine 2015;372:1009–1018.

4. Goyal M, Demchuk AM, Menon BK, Eesa M, Rempel JL, Thornton J, Roy D, Jovin TG, Willinsky RA, Sapkota BL, et al. Randomized assessment of rapid endovascular treatment of ischemic stroke. New England Journal of Medicine 2015;372:1019–1030.

5. Goyal M, Menon BK, van Zwam WH, Dippel DW, Mitchell PJ, Demchuk AM, Dávalos A, Majoie CB, van der Lugt A, de Miquel MA, et al. Endovascular thrombectomy after large-vessel ischaemic stroke: a meta-analysis of individual patient data from five randomised trials. Lancet 2016;387:1723–1731.

6. Jovin TG, Chamorro A, Cobo E, de Miquel MA, Molina CA, Rovira A, San Román L, Serena J, Abilleira S, Ribó M, et al. Thrombectomy within 8 hours after symptom onset in ischemic stroke. New England Journal of Medicine 2015;372:2296–2306.

7. Nogueira RG, Jadhav AP, Haussen DC, Bonafe A, Budzik RF, Bhuva P, Yavagal DR, Ribo M, Cognard C, Hanel RA, et al. Thrombectomy 6 to 24 hours after stroke with a mismatch between deficit and infarct. New England Journal of Medicine 2018;378:11–21.

8. Saver JL, Goyal M, Bonafe A, Diener HC, Levy EI, Pereira VM, Albers GW, Cognard C, Cohen DJ, Hacke W, et al. Stent-retriever thrombectomy after intravenous t-PA vs. t- PA alone in stroke. New England Journal of Medicine 2015;372:2285–2295.

9. Goyal M, Ospel JM, Menon B, Almekhlafi M, Jayaraman M, Fiehler J, Psychogios M, Chapot R, van der Lugt A, Liu J, et al. Challenging the ischemic core concept in acute ischemic stroke imaging. Stroke 2020;51:3147–3155.

10. Morita N, Harada M, Uno M, Matsubara S, Matsuda T, Nagahiro S, Nishitani H. Ischemic findings of T2*-weighted 3-tesla MRI in acute stroke patients. Cerebrovascular Diseases 2008;26:367–375.

11. Terasawa Y, Yamamoto N, Morigaki R, Fujita K, Izumi Y, Satomi J, Harada M, Nagahiro S, Kaji R. Brush sign on 3-T T2*-weighted MRI as a potential predictor of hemorrhagic transformation after tissue plasminogen activator therapy. Stroke 2014;45:274–276.

12. Purushotham A, Campbell BC, Straka M, Mlynash M, Olivot JM, Bammer R, Kemp SM, Albers GW, Lansberg MG. Apparent diffusion coefficient threshold for delineation of ischemic core. International Journal of Stroke 2015;10:348–353.

13. Yoshimoto T, Inoue M, Yamagami H, Fujita K, Tanaka K, Ando D, Sonoda K, Kamogawa N, Koga M, Ihara M, et al. Use of diffusion-weighted imaging-Alberta stroke program early computed tomography score (DWI-ASPECTS) and ischemic core volume to determine the malignant profile in acute stroke. Journal of the American Heart Association 2019;8:e012558.

14. Inoue M, Mlynash M, Christensen S, Wheeler HM, Straka M, Tipirneni A, Kemp SM, Zaharchuk G, Olivot JM, Bammer R, et al. Early diffusion-weighted imaging reversal after endovascular reperfusion is typically transient in patients imaged 3 to 6 hours after onset. Stroke 2014;45:1024–1028.

15. Li F, Liu KF, Silva MD, Omae T, Sotak CH, Fenstermacher JD, Fisher M, Hsu CY, Lin W. Transient and permanent resolution of ischemic lesions on diffusion-weighted imaging after brief periods of focal ischemia in rats: correlation with histopathology. Stroke 2000;31:946–954.

16. Li Y, Wang T, Zhang T, Lin Z, Li Y, Guo R, Zhao Y, Meng Z, Liu J, Yu X, et al. Fast high-resolution metabolic imaging of acute stroke with 3D magnetic resonance spectroscopy. Brain 2020;143:3225–3233.

17. Labeyrie MA, Turc G, Hess A, Hervo P, Mas JL, Meder JF, Baron JC, Touzé E, Oppenheim C. Diffusion lesion reversal after thrombolysis: a MR correlate of early neurological improvement. Stroke 2012;43:2986–2991.

18. Tisserand M, Malherbe C, Turc G, Legrand L, Edjlali M, Labeyrie MA, Seners P, Mas JL, Méder JF, Baron JC, et al. Is white matter more prone to diffusion lesion reversal after thrombolysis? Stroke 2014;45:1167–1169.

